# TopBrain Segmentation Challenge for Whole Brain Vessel Anatomy

**DOI:** 10.64898/2026.05.28.26354312

**Authors:** Kaiyuan Yang, Pengcheng Shi, Houjing Huang, Fabio Musio, Hakim Baazaoui, Orhun Utku Aydin, Adam Hilbert, Rachika E. Hamadache, Cansu Yalcin, Minghui Zhang, Daniele Falcetta, Ezequiel de la Rosa, Suprosanna Shit, Chinmay Prabhakar, Bastian Wittmann, Maximilian R. Rokuss, Yannick Kirchhoff, Rami Al-Maskari, Luciano Höher, Norman Juchler, Adrià Casamitjana, Jon Cleary, Anton Schmick, Philipp Baumgartner, Julian Deseö, Olafs Vandans, Dahye Lee, Kwanseok Oh, Dominic LaBella, Moona Mazher, Steven A. Niederer, Abdul Qayyum, Yaoyu Liu, Junqiang Chen, Wooseung Kim, Napasara Asawalertsak, Minjae Kim, Dongho Shin, Sung-Hong Park, Shunsuke Kikuchi, Yaqing Zhang, Jialu Liu, Yue Cui, Yuchen Qiu, Anouk Verschuur, Jiaxin Zhang, Irene van der Schaaf, Ruisheng Su, Chantal M.W. Tax, Yosuke Yamagishi, Nishta Letchumanan, Shouhei Hanaoka, Jesús González, Riccardo Tiberi, Clara Lisazo, Valeriia Abramova, Uma Maria Lal-Trehan Estrada, Agustin Cartaya Lathulerie, Micaela Rivas Díaz, Arnau Oliver, Menghan Zhang, Zhiqiang Bai, Klaus Maier-Hein, Yun Gu, Xavier Lladó, Maria A. Zuluaga, Dietmar Frey, Johannes C. Paetzold, Sven Hirsch, Susanne Wegener, Yihui Ma, Bjoern Menze

**Author notes:** Y.M. and B.M. are co-corresponding authors (Yihui Ma), (Bjoern Menze). K.Y., P.S., and H.H. contributed equally. Participant of the challenge, ordered alphabetically by team name.

## Abstract

We present the TopBrain 2025 Challenge, the first benchmark for fine-grained multi-class segmentation of the whole brain vasculature in both computed tomography angiography (CTA) and magnetic resonance angiography (MRA). Building on the TopCoW challenge, TopBrain scales vessel annotation from the Circle of Willis to the entire brain, introducing a dataset of 90 annotated volumes across 48 landmark vessel classes spanning arterial and venous systems, of which 50 training volumes are publicly released. Vessel definitions were consolidated from established neuroanatomical references into a unified annotation scheme, and vessel caliber measurements along the centerline are reported for the first time across the whole brain vascular anatomy. To address the unique challenges of multiclass brain vessel segmentation, we propose an evaluation framework that accounts for detection in segmentation performance, assesses anatomical plausibility, and introduces novel contamination metrics that characterize inter-class prediction errors. Fifteen teams from over 220 registered participants submitted algorithms to the benchmark. The top-performing teams built on nnUNet with principled system design choices, achieving around 80% Dice scores, near-zero invalid neighbor counts, over 60% F1 scores for side-road vessels, and below 18% fore-ground contamination ratio. Larger vessels are easier to segment, while smaller and more complex vessels remain the true bottleneck. The annotated datasets and podium-finish algorithms are made publicly available on Zenodo.

## 1. Introduction

The human brain vasculature is an intricate network of arteries and veins characterized by complex morphology and considerable inter-subject variability. Accurate segmentation of these structures is essential for understanding brain perfusion and diagnosing vascular pathologies. However, this task is non-trivial; the vasculature consists of an intertwined, tortuous architecture where vessels of varying calibers are packed in close proximity, often in narrow and spatially irregular confines of fissures, sulci, bony canals, and subarachnoid cisterns. Resolving vessel-class ambiguity further requires understanding both the spatial relationships between vessels and the surrounding anatomical context, often with substantial anatomical variations.

Despite recent advances, studies on brain vascular segmentation remain limited by several methodological shortcomings. First, many prior works do not explicitly describe their annotation protocols or vessel-label definitions, particularly in the presence of anatomical variants. Because there is no consensus on the anatomical boundaries of many vessel labels in the brain, the absence of explicit label definitions hinders reproducibility and cross-study comparison. In addition, most existing datasets and annotations are not publicly released, which prevents independent validation of anatomy and the development of standardized benchmarks.

Second, existing studies had insufficient label granularity and anatomical coverage, typically focusing on large, high-contrast vessels while omitting smaller or anatomically complex structures. Although prior research has explored multi-class cerebrovascular segmentation, the number of annotated vessel classes in existing works was restricted to below 20 and included only arteries [1, 2, 3, 4, 5, 6]. Such limited classification schemes lack the anatomical granularity and scale to differentiate the full brain vascular anatomy.

Third, current evaluation strategies are insufficient to address problems of multiclass brain vascular segmentation. The brain vasculature exhibits substantial anatomical variation, where specific vessels may be absent or aplastic; consequently, segmentation performance is heavily confounded by detection performance, a distinction often ignored by standard metrics. Furthermore, the close proximity of vessels traversing within the winding narrow confines leads to significant inter-class confusion between nearby vessels, whereas conventional metrics evaluate each class independently. Existing evaluation schemes also fail to account for anatomical plausibility, such as valid neighborhood relationships between classes. Specific evaluation metrics are thus needed to capture the structural and topological errors unique to multiclass brain vascular segmentation.

To bridge these research gaps, we present the TopBrain 2025 Challenge, which released a publicly available annotated dataset and collected crowd-sourced algorithms for segmentation performance benchmark. TopBrain challenge serves as an extension of the previous TopCoW challenge [7], scaling the annotated vessel anatomy from the Circle of Willis (CoW) to the whole brain area. The resulting TopBrain dataset represents the first fine-grained 42-class multiclass segmentation dataset encompassing both the arterial and venous systems of the brain. Annotations were performed on two widely used non-invasive angiographic modalities, computed tomography angiography (CTA) and magnetic resonance angiography (MRA). By surveying relevant anatomy literature, we unified vessel definitions into an annotation scheme that is scalable, robust, and grounded in clinical utility. As a by-product of our annotation, we report vessel caliber measurements along the centerline of all annotated vessels. To our knowledge, this is the first study to systematically quantify vessel caliber over the whole brain vessel anatomy. We propose an evaluation framework tailored for brain vessel anatomy segmentation that decouples detection performance, accounts for anatomical plausibility and interclass contamination. For transparency and to support continued development, we have made our annotated dataset and podium-finish algorithm submissions publicly available on Zenodo^2^.

## 2. Study Design

For ease of reference, the anatomical abbreviations used in this paper are listed in the Appendix A.

### 2.1. TopBrain Dataset

The TopBrain dataset comprises 90 volumes, partitioned into a training set of 50 images and a test set of 40 images, with each set containing an equal distribution of MRA and CTA modalities. These images constitute a subset of the previously released TopCoW challenge datasets. In addition to the exclusion criteria established in TopCoW, the TopBrain cohort further excluded cases with bihemispheric A2 or A3 variants to maintain label consistency for the ACA vessels. Given the stroke-center origin of the TopCoW cohort, the TopBrain dataset includes numerous cases characterized by large vessel occlusions (LVO) and severe stenosis across various vascular regions.

### 2.2. Data Annotation Workflow

Vessel labeling was performed using the virtual-reality (VR) annotation workflow established in the TopCoW challenge. The vessel annotation protocol was designed by a senior neurosurgeon (Y.M., over 10 years of experience). Voxel-level annotations were performed by a doctoral researcher (K.Y.) after training by Y.M. who used the initial four patients to instruct the annotator on anatomical knowledge and the annotation protocol. Subsequent images were pre-annotated by a segmentation model progressively refined with increasing amounts of training data via an iterative “human-in-the-loop” approach. These pre-labels were then manually corrected and verified at the voxel level by K.Y. Any cases involving complex anatomy or labeling uncertainty were further verified and adjudicated by Y.M.

### 2.3. Atlas of Annotated Vessels

Inspired by the visual style of metro traffic maps, we illustrate the anatomical atlas of the annotated vessels in Fig. 1a. Using the analogy of metro lines, TopBrain defines seven major “vessel lines”: the VB, PCA, ICA, ACA, MCA, ECA, and Vein lines. We annotated 42 vessel classes in the MRA modality and 40 classes in the CTA modality, of which 34 classes are common across both modalities. The ECA line is unique to MRA, whereas the Vein line is unique to CTA.

**Fig. 1.**
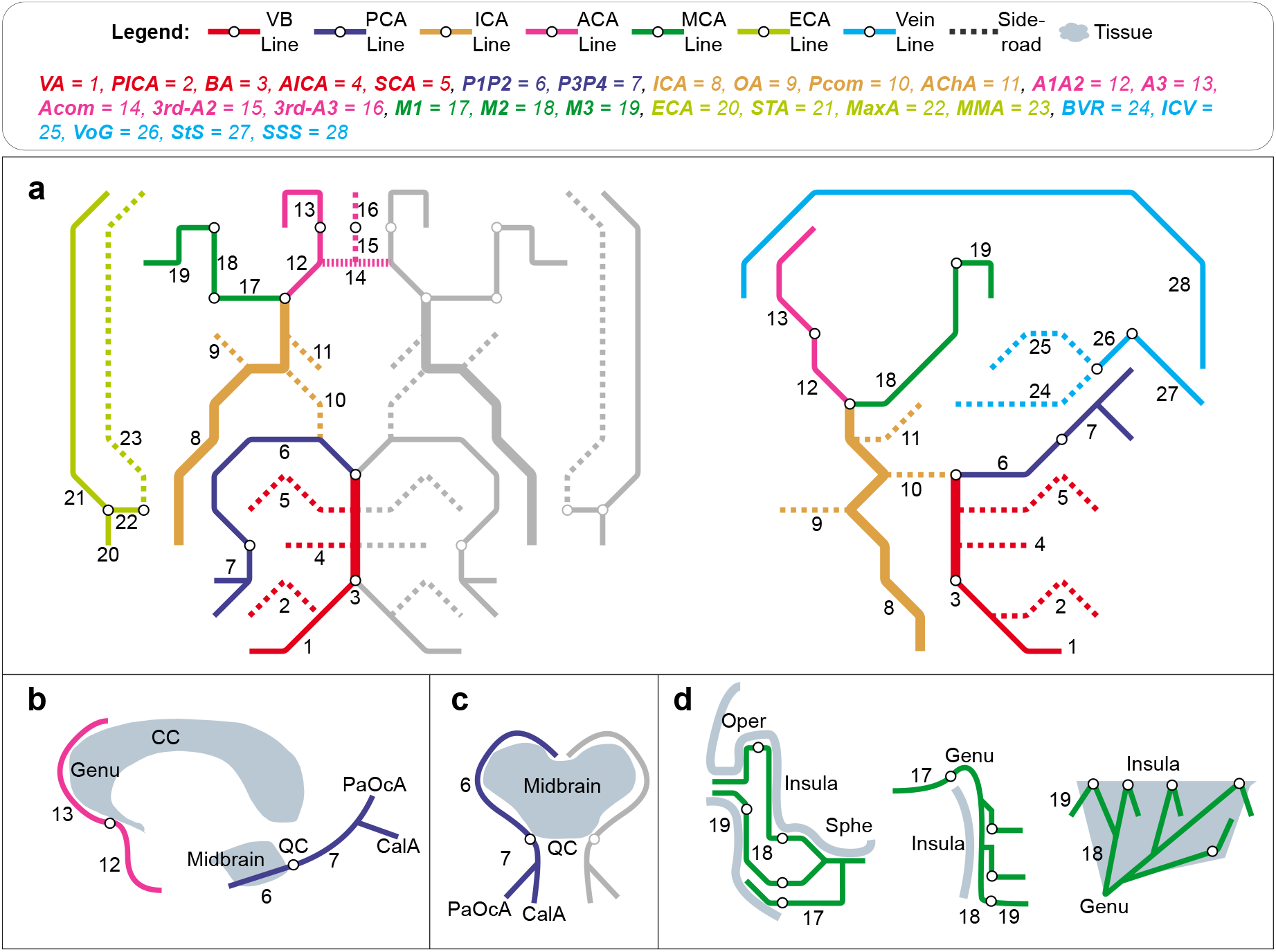
TopBrain anatomy atlas in the style of metro maps. (a) Overview of all the anatomy labels in the TopBrain dataset. Left shows the submentovertex view and right shows the anterior-posterior view. (b) Sagittal view of the ACA and PCA lines in relationship to the surrounding brain tissues. (c) Axial view of the PCA line and nearby brain tissues. (d) From left to right, coronal, axial, and sagittal views of the relationship between MCA line and nearby brain tissues.

Using an additional transportation analogy, vessels were further categorized into “highway” and “side-road” vessels. Highway vessels correspond to major vessels for each vessel line with relatively large caliber and prominent blood supply. In contrast, side-road vessels are typically thinner branches originating from the highway vessels and may be absent or difficult to visualize in angiographic imaging because of weak signal intensity or aplasia. Fig. 1b to 1d illustrates the anatomical context to segment the ACA, PCA, and MCA lines.

Table 1 lists the anatomical descriptions and definitions for each annotated vessel in terms of origin, course, and termination, along with excluded variants, measured caliber, and projected clinical interest. The vessel caliber measurement followed the method described in [8]. The median diameter reported in Table 1 was aggregated from the training set, using the CTA modality when available and the MRA modality otherwise. Note that the caliber measurements in the results section were obtained separately from the CTA and MRA test sets for each vessel class. The clinical interest ratings reflect the expert judgment of a senior neurosurgeon (Y.M.) on a scale of 0 to 4, separately for stenosis/occlusion and intracranial aneurysm relevance.

**Table 1.**
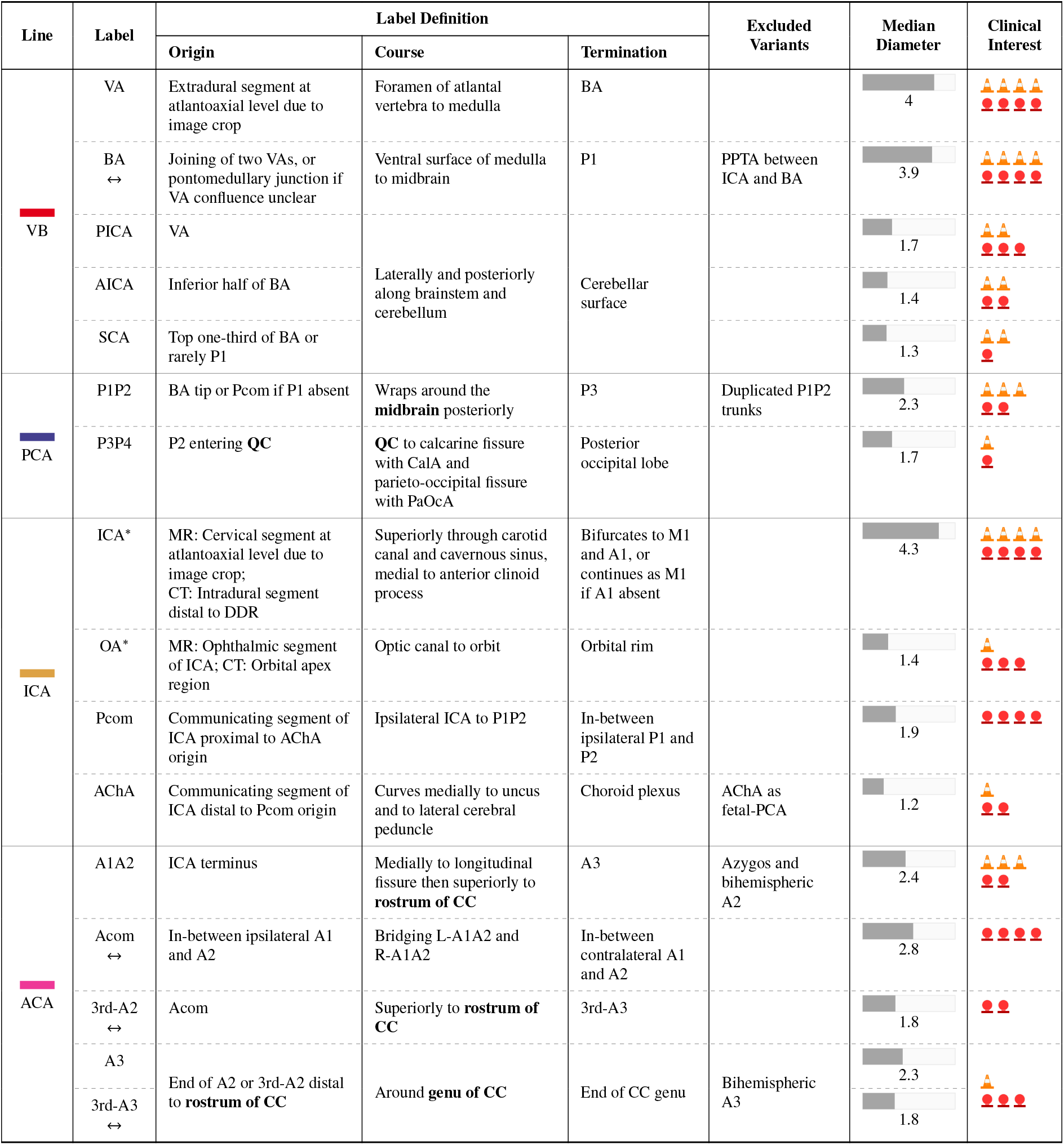
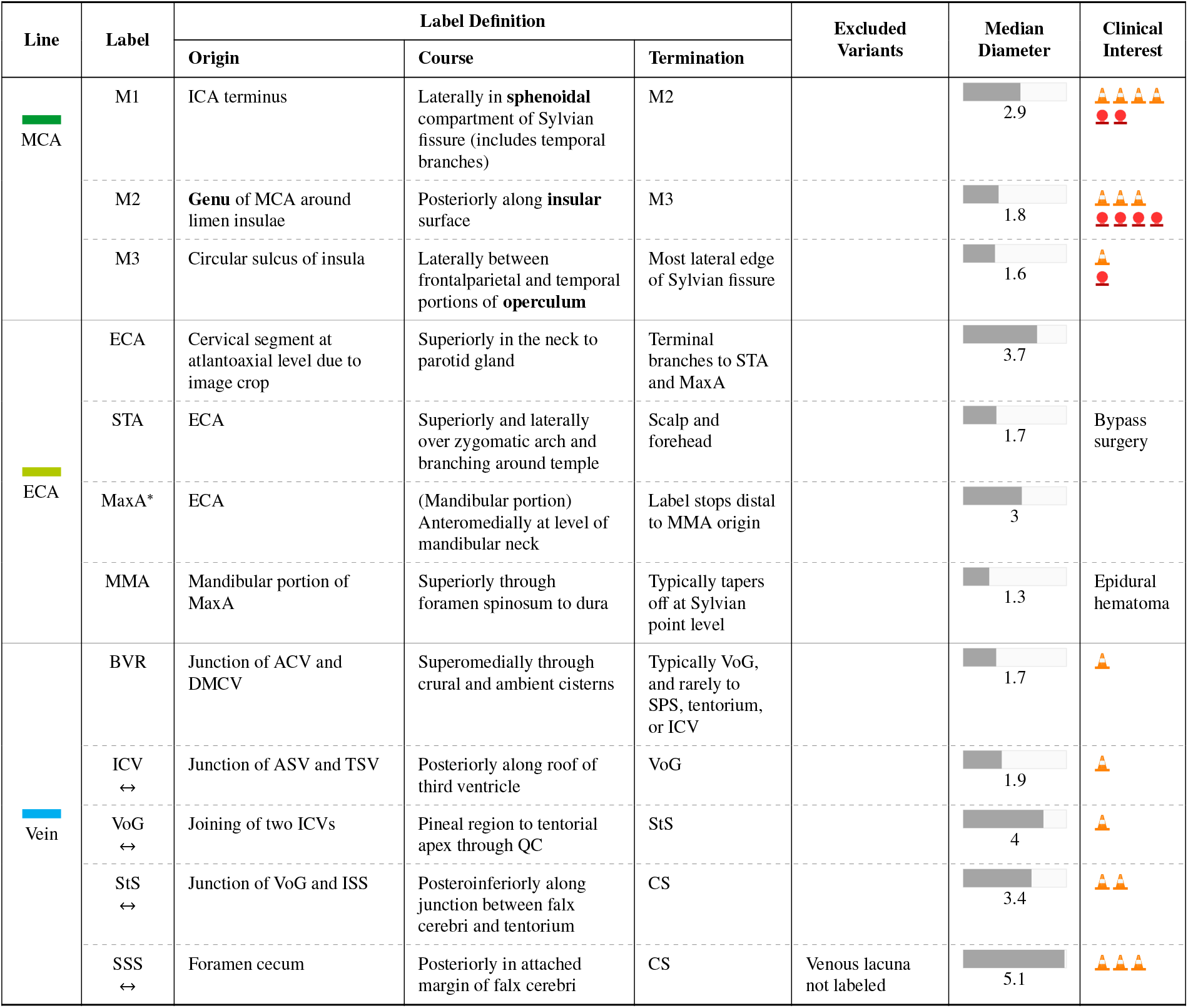
Annotated vessels in the TopBrain dataset. List of abbreviations can be found in Appendix A. In the ‘Label’ column, the symbol ↔denotes vessels assigned a single label without differentiating laterality, and the asterisk ^*^ indicates vessels that were partially annotated in at least one modality. The ‘Label Definition’ column specifies the segmentation boundaries, with selected anatomical landmarks shown in bold. In the ‘Median Diameter’ column, measurements were obtained from the training set using the CTA modality when available and the MRA modality otherwise (units: millimeters). In the ‘Clinical Interest’ column, 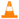 represents the relevance level for stenosis/occlusion, and 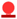 represents the relevance level for intracranial aneurysm (on a scale of 0 to 4).

### 2.4. Consolidation of Vessel Definitions

The vessel definitions used in this study were synthesized from established neuroanatomical and neuroradiological references. Osborn’s Brain [9] and Radiopaedia (https://radiopaedia.org) served as the primary references for general anatomical framework. For vascular systems with greater anatomical variability or labeling ambiguity, additional literature was consulted, including works addressing ACA segments [10, 11, 12, 13, 14, 15, 16], MCA segments [17, 18, 19, 20, 21, 22, 23], and venous anatomy [24, 25, 26, 27, 28].

It is known that vessel definitions vary among practitioners and even among established references. Importantly, rather than strictly adhering to any single source, we unified and consolidated definitions across references. The result is an annotation scheme optimized for generalizable and practical use in large-scale vessel annotation.

### 2.5. Challenge Structure

The 2025 challenge (https://topbrain2025.grand-challenge.org) consisted of two modality tracks, MRA and CTA, for algorithm submissions. All algorithms were required to be fully automatic and submitted as Docker containers. Inference was performed in the cloud on the submission platform, which provided an NVIDIA T4 GPU with 16 GB of GPU memory and 31 GB of main memory. Each algorithm had a maximum runtime of 15 minutes per test case. For each track’s hidden test set, teams were allowed only one container submission to prevent tuning to the test set.

### 2.6. Evaluation of Algorithms

The segmentation predictions were evaluated using a set of 10 metrics, grouped into three categories: standard segmentation metrics, contamination metrics, and tailored vascular metrics.

#### Standard Segmentation Metrics

The four standard segmentation metrics are the Dice similarity coefficient (Dice score), centerline Dice (clDice) [29], Hausdorff distance at the 95th percentile (HD95), and connected component (zero-th Betti number) error.

For each test case, we compute the metric for every class in the union of classes present in the ground truth and the prediction (i.e., excluding classes absent from both). This ensures that the evaluation accounts for false positive and false negative detection failures. Finally, we report the per-case class-average, calculated as the mean of the class-average scores across the entire test set.

#### Contamination Metrics

To characterize inter-class prediction errors, we propose four contamination metrics: foreground contamination (FGC) ratio, count of FGC sources, count of background contamination (BGC) voxels, and count of BGC sources, as illustrated in Fig. 2. Each ground-truth (GT) voxel is first assigned to one of four types: *foreground* (FG), *interface* (boundary between two FG classes), *background* (BG), or *surface* (boundary between BG and FG). To account for uncertainty in boundary delineation, the *interface* thickness was configured to three voxels per class (extending into both adjacent FG regions), and the *surface* thickness was set to five voxels (mono-directionally shrinking the BG region). The FGC ratio is the percentage of incorrectly predicted FG voxels, and the count of FGC sources is the number of contributing classes, both averaged over all classes in the GT. The counts of BGC voxels and BGC sources report the number of incorrectly predicted BG voxels and their contributing classes, respectively.

**Fig. 2.**
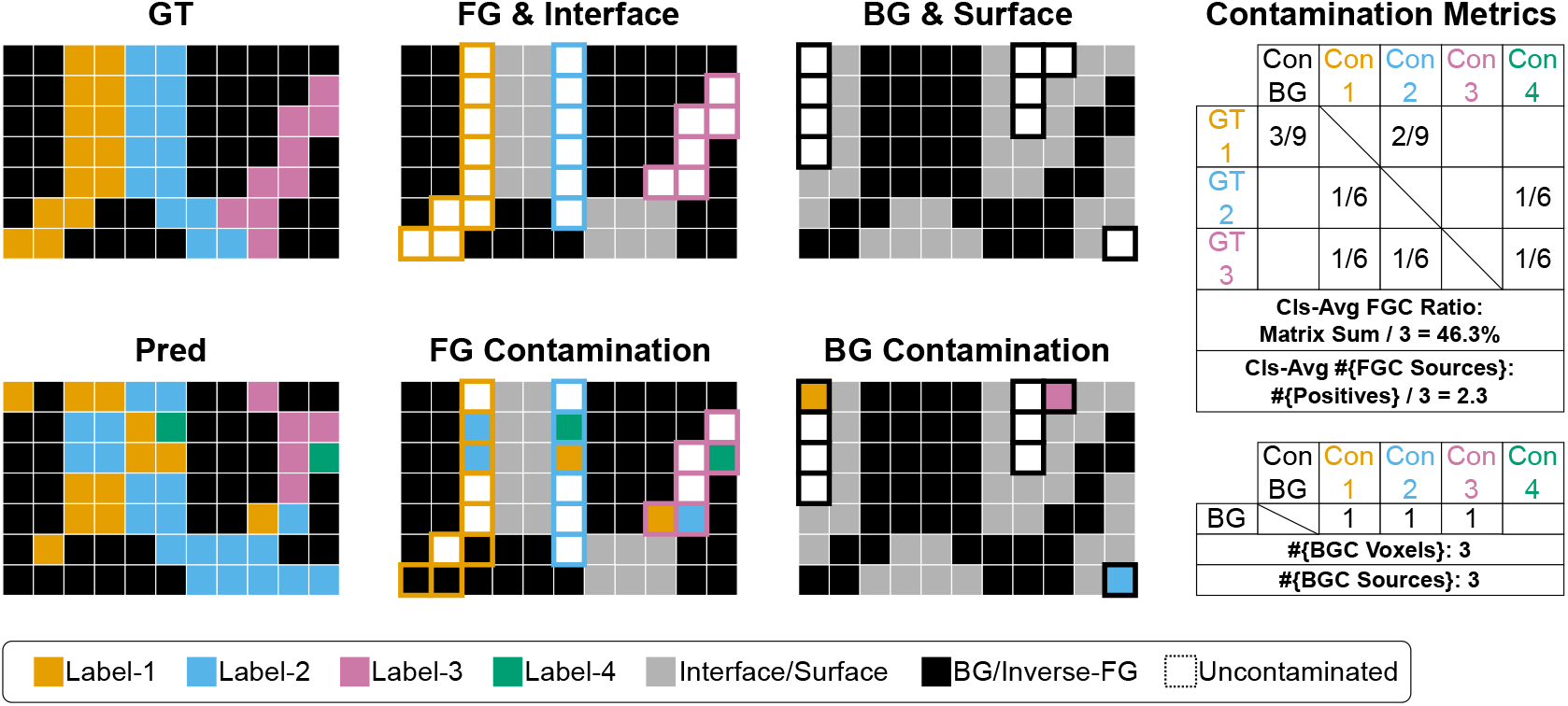
Toy example of the proposed contamination metrics. We illustrate the metric computation using an 11×7 pixel grid featuring four vessel labels. The ground-truth contains three class labels, while the prediction includes various contamination scenarios from the three labels and a false positive (Label-4). A uniform 1-pixel thickness is used for *surface* and *interface* for simplicity. Abbreviations: background (BG), background contamination (BGC), class-average (Cls-Avg), contaminant (Con), foreground (FG), foreground contamination (FGC), ground-truth (GT), and prediction (Pred).

#### Tailored Vascular Metrics

Two metrics were tailored for the vascular context of the task: the invalid neighbor count and the F1 score of side-road vessels.

The invalid neighbor count quantifies anatomical implausibility in the predicted segmentation. For each class in the anatomy atlas, a list of valid neighboring classes is precompiled from the training set, defined as any class pair that was ever observed to be adjacent. During evaluation, for each predicted class, we count the number of neighboring classes not belonging to the pre-compiled valid neighbor list. The metric is reported as the per-case class-average, computed over all classes present in the prediction.

The F1 score for detection of side-road vessels is reported as the macro per-class average. A vessel instance is considered a true positive if the intersection over union between the predicted and ground-truth masks is at least 25%. The distribution of side-road vessels from the training and test sets is provided in Appendix B.

#### Ranking

Algorithm ranking for the leaderboards was based on the four standard segmentation metrics and the two tailored vascular metrics. The final leaderboard rank was determined by averaging the rank positions across all six metrics. The contamination metrics were introduced after the leaderboard ranking procedure and were therefore used only for the additional analyses presented in this paper.

## 3. Results

### 3.1. Algorithm Characteristics

In the 2025 challenge, 15 teams out of over 220 registered participants from six continents submitted algorithms to the benchmark. Of these, nine teams developed algorithms for both MRA and CTA modalities.

Interestingly, several design choices converged among participants as shown in Table 2. The nnUNet [30] remained a favorite framework, utilized by 11 out of the 15 teams. Five teams combined both binary and multiclass segmentation models, leveraging the former to refine the results of the latter. Three teams grouped the brain vessels into anatomically themed sub-groups to tackle segmentation by sub-region. Three teams trained their models with mixed modalities. Five teams employed some form of pretraining for one of the modalities; these strategies included fine-tuning self-supervised learning (SSL) models, adapting vessel foundation models, or initializing weights from previously trained vessel models. Additionally, four teams conducted connected component removal in the post-processing. Summary descriptions for each team’s method are provided in Appendix C.

**Table 2.**
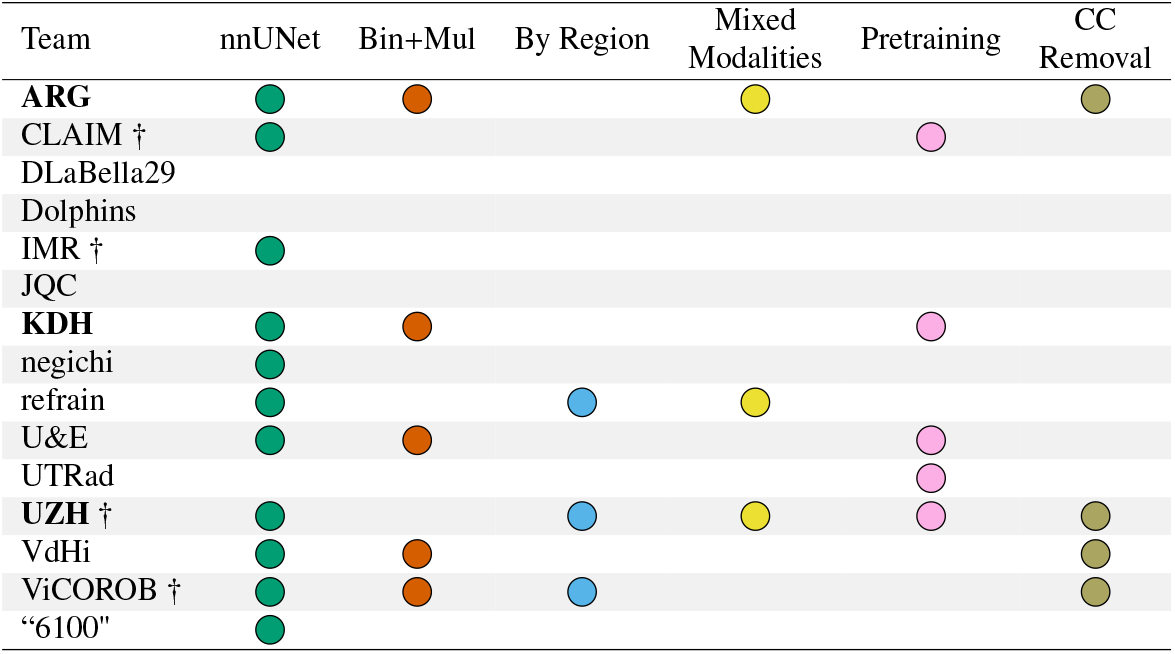
Common design strategies used by the 2025 teams. Teams are ordered alphabetically by team names. Highlighted cells indicate the use of a given strategy. indicates teams that include at least one organizing committee member. Teams with podium finish for either modalities are highlighted in bold. Abbreviations: binary (Bin) and multiclass (Mul) segmentation; connected component (CC).

### 3.2. Overall Quantitative Results

Table 3 shows the results of the 2025 multiclass segmentation task for CTA and MRA modalities. The same three teams achieved top-3 finish for both modalities. We highlight a few observations for the best-performing teams. The per-case class-average Dice reached around 80%. The invalid neighbor count was close to zero. Side-road vessels were moderately detected with F1 scores around 68% for CTA and 85% for MRA. Inter-class contamination metrics indicated that around 18% of the foreground voxels were contaminated by incorrect class predictions, with the source limited to approximately 1.2 to 1.3 classes on average. The background was contaminated by 500–1000 voxels, which is roughly the size of a prominent Acom vessel, originating from 5–6 vessel classes.

**Table 3.**
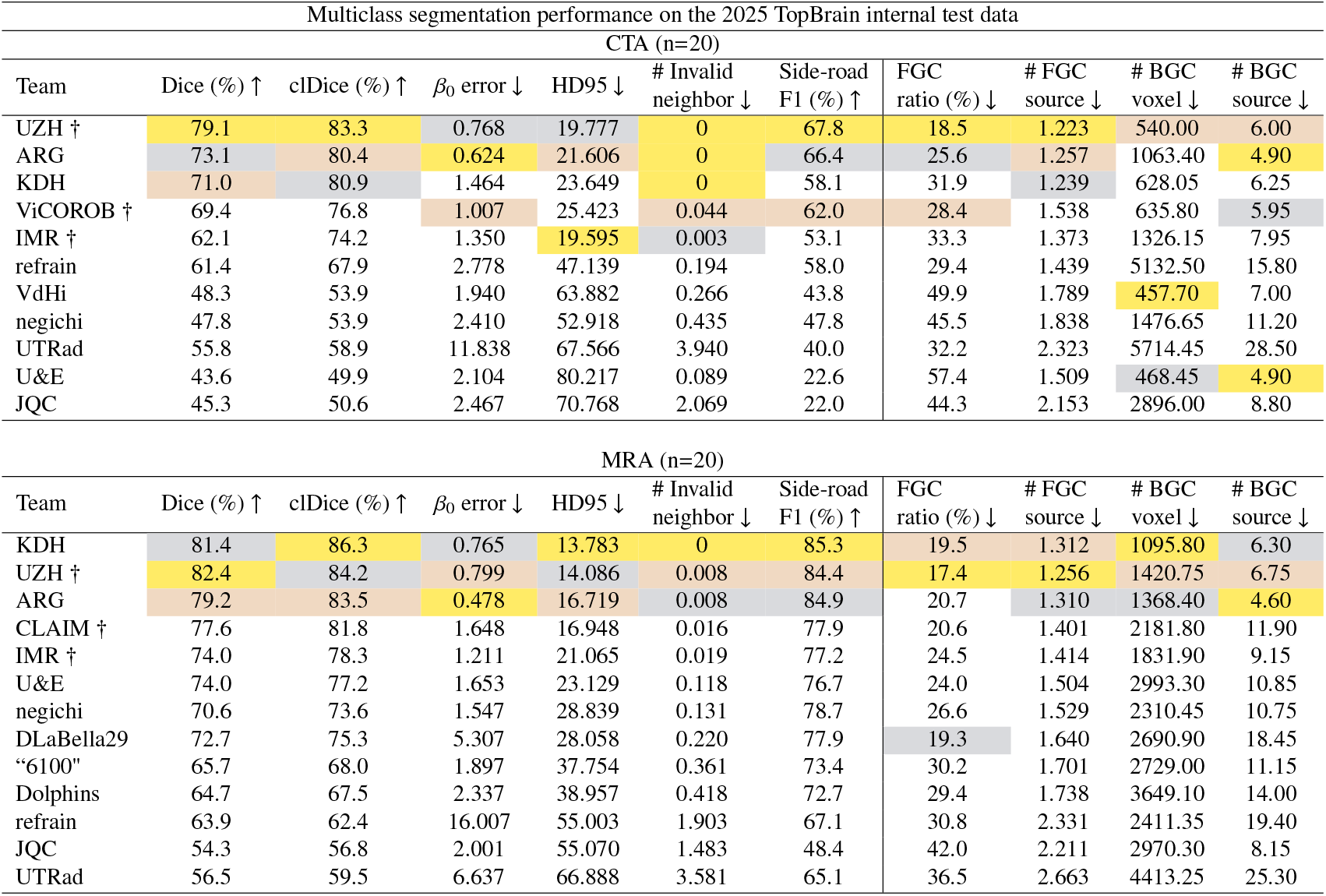
Results in mean values of the multiclass segmentation task from 2025 on the TopBrain internal test set in terms of Dice, centerline Dice (clDice), connected component number (0-th Betti number *β*_0_) error, Hausdorff Distance 95% Percentile (HD95), count (#) of invalid neighbors, F1 score for detection of side-road vessels, foreground contamination (FGC) ratio, count of FGC sources, count of background contamination (BGC) voxels, and count of BGC sources. All metrics represent the per-case class-average, except for the side-road F1 score, which is a global per-class average. The arrow indicates the favorable direction. The top three values for each metric are marked in gold, silver, and bronze colors. The ‘Team’ column is ordered by ranking from the challenge leaderboard. indicates teams that include at least one organizing committee member.

Performance on MRA was generally higher than on CTA for all nine teams that participated in both modalities. For the 34 labels common to both modalities, the Dice scores were 8–15 points higher and HD95 scores were half as large in MRA than in CTA for the best two teams, suggesting that CTA is a harder modality to segment.

### 3.3. Vessel Caliber vs Segmentation

Fig. 3 plots the diameter of each vessel class against its segmentation performance in terms of Dice and side-road F1 scores. Larger vessel caliber is strongly associated with reliable segmentation performance: vessels with median diameters of 2 mm or greater almost consistently achieve Dice and F1 scores above roughly 70%, while all vessels larger than 3 mm achieve Dice scores above 86%. Nevertheless, strong performance is not exclusive to large vessels, as several vessels smaller than 2 mm also achieve high Dice and F1 scores.

**Fig. 3.**
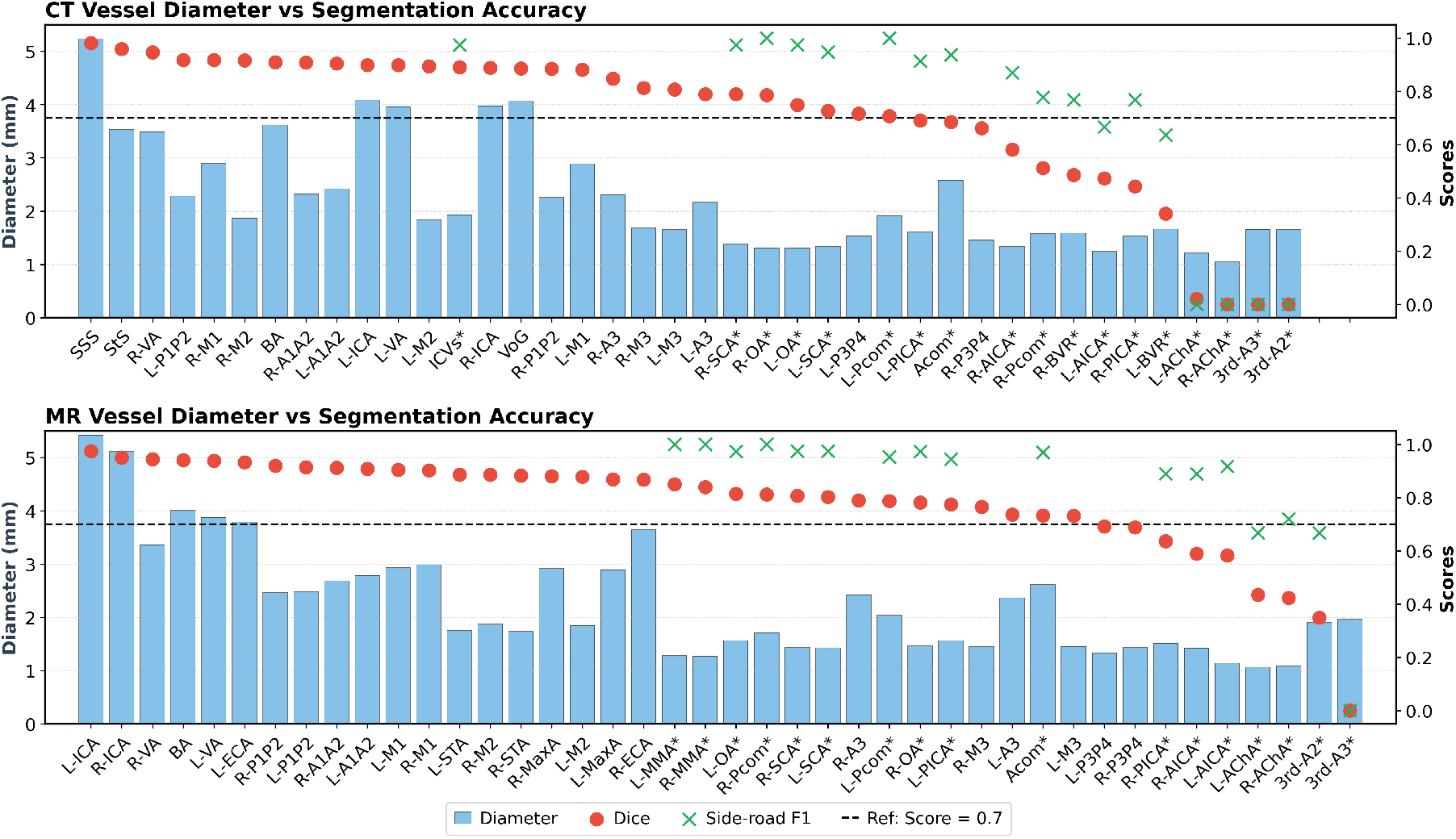
Vessel diameter and segmentation accuracy. CT (top) and MR (bottom) vessel diameter versus average Dice and side-road F1 scores. The vessels are sorted in descending Dice score order. Segmentation scores are from the top team from each modality: team ‘UZH’ for CT and team ‘KDH’ for MR.

Side-road vessels generally have calibers below 2.6 mm. A positive correlation between their F1 and Dice scores can also be observed, indicating that detection failure is a major factor affecting segmentation performance for side-road vessels. Among these, L-/R-AChA, 3rd-A2, and 3rd-A3 stand out as particularly challenging vessels for detection and segmentation, likely due in part to their low prevalence in the dataset (Table B.4).

### 3.4. Hard Situations for Segmentation

Vascular structures do not present uniform levels of segmentation difficulty. Certain anatomical regions exhibit intricate topological relationships and require strict adherence to anatomical definitions of origin and course to resolve class ambiguity. We identify four representative “hard situations” that highlight these challenges in Fig. 4. Notably, these challenges are not limited to automated segmentation. The same anatomical complexities also complicate manual annotation, as they pose difficulties even for expert clinical verification.

**Fig. 4.**
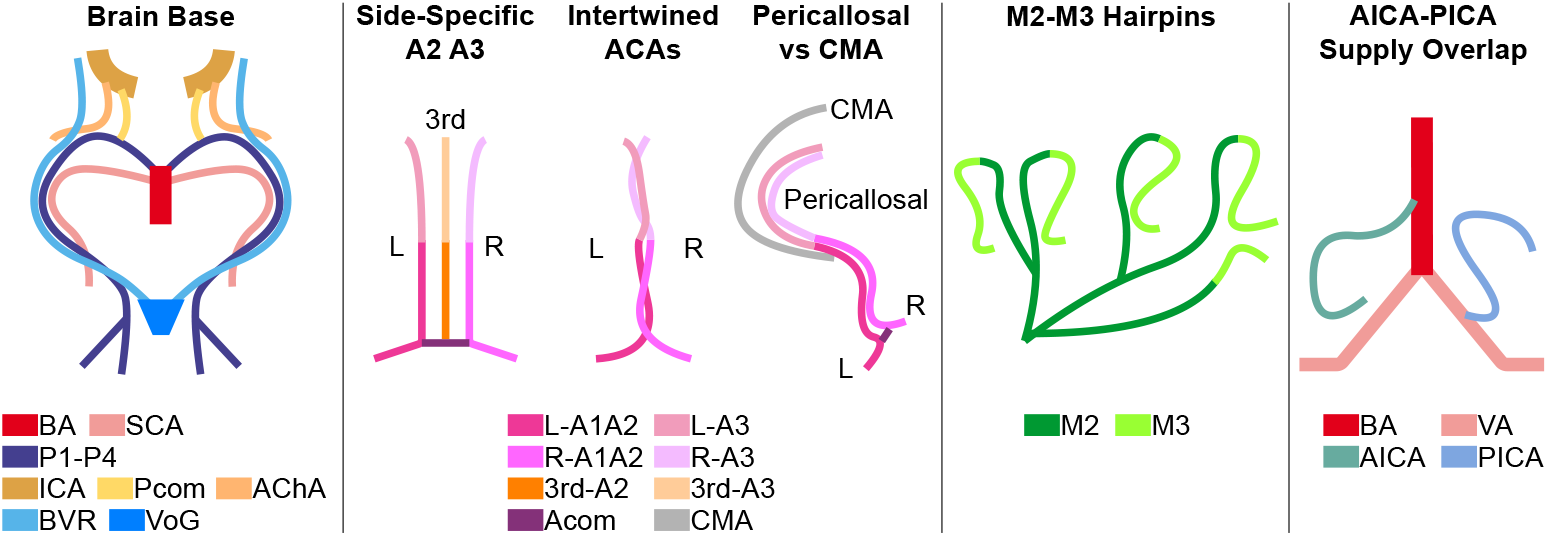
Hard situations for segmentation. Left panel: arteries and veins run in parallel and close proximity, while adhering to increasing z-axis order, at the base of the brain. Second panel: difficulties for the ACA line. Third panel: difficulties due to the M2-M3 hairpins for the MCA line. Fourth panel: difficulties due to origin and course for the AICA-PICA vessels in the VB line.

1. *Brain base*. At the base of the brain, arteries and veins traverse narrow cisternal spaces, resulting in dense spatial overlap while maintaining a strict inferior-superior (z-axis) spatial relationship. For example, the origin of Pcom is inferior to that of AChA on the ICA. In the crural cistern, AChA and BVR often run adjacent to each other. In the ambient cistern, these vessels typically follow a z-axis order from inferior to superior: SCA, PCA, and BVR.
2. *ACAs*. Within the interhemispheric fissure, the ACAs must be tracked with strict preservation of laterality and segmental continuity (e.g., L-A2 to L-A3, R-A2 to R-A3, and 3rd-A2 to 3rd-A3). However, these vessels are frequently intertwined, making it difficult to disentangle individual A2-A3 courses. In addition, the CMA, which courses farther from the genu of CC, may be confused with the pericallosal A3 segment, which runs along the CC genu.
3. *M2-M3 hairpin*. At the circular sulcus, each M2 segment typically gives rise to its corresponding M3 segment via a hairpin turn. Differentiating between M2 and M3 in these loops is difficult, as it depends on correctly interpreting vessel direction relative to the insular cortex.
4. *AICA-PICA supply overlap*. Despite their often similar supply territories, the distinction between AICA and PICA is defined primarily by vessel origin: AICA arises from the inferior half of BA, whereas PICA originates from VA. This distinction becomes particularly challenging in cases of AICA-PICA common trunk variants, where identification must rely exclusively on identifying the origin while disregarding distal course.

Fig. 5 shows the qualitative results on the four hard situations from each modality. The MRA case has a class-average Dice of 73.0%, 0 invalid neighbors, and 73.3% side-road F1 score. The CTA case has a class-average Dice of 78.6%, 0 invalid neighbors, and 91.7% side-road F1 score. False-positive and false-negative detections can be seen in brain-base, ACA, and VB regions for the MRA case, contributing to its reduced F1 score. Greater inter-class confusion in MRA also leads to elevated foreground contamination scores. The CTA case exhibits more over-segmentation errors in the background, as reflected in the higher number of BGC metrics.

**Fig. 5.**
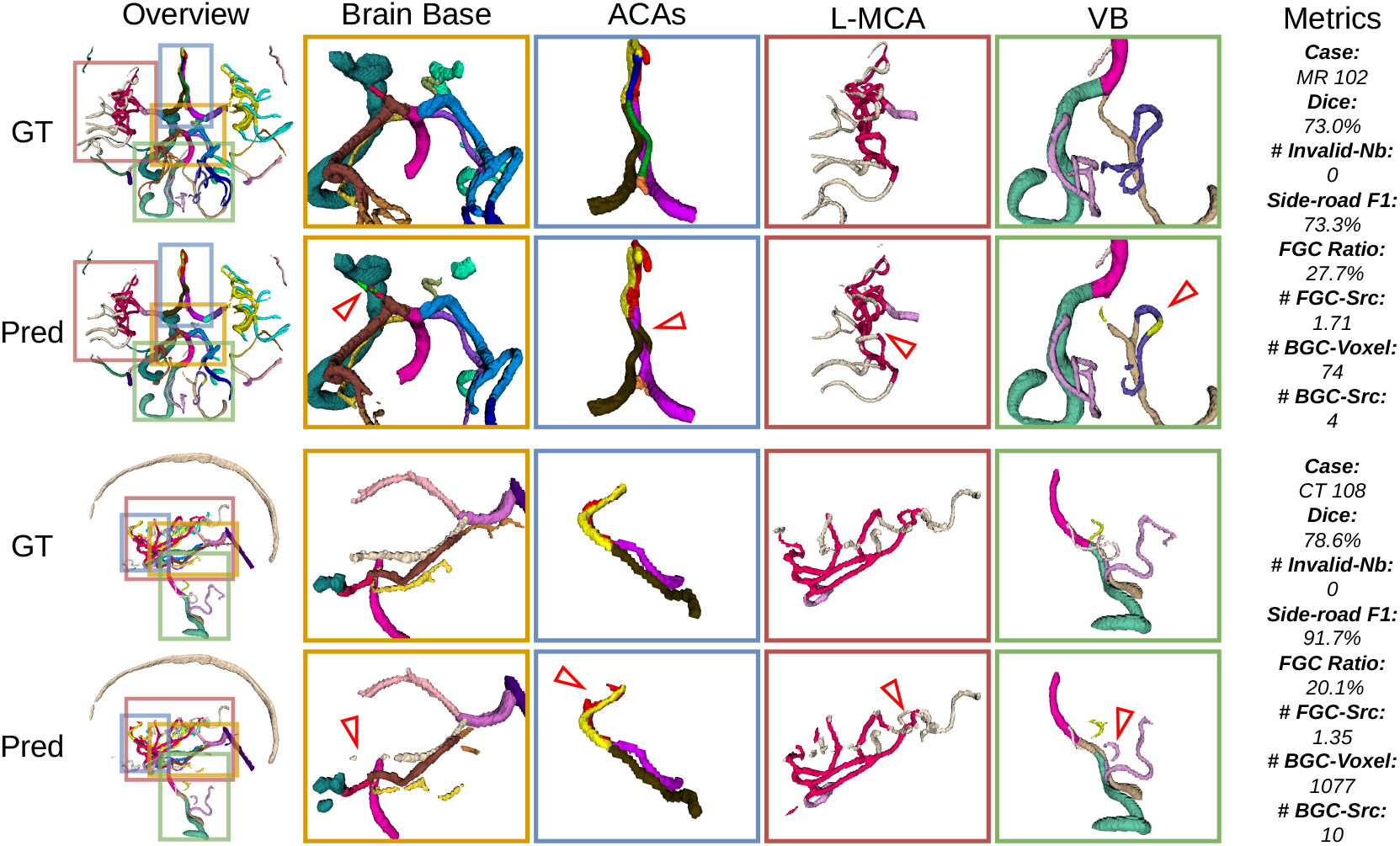
Qualitative segmentation results with case-specific metrics. Two cases are shown: one MRA and one CTA. Predictions (Pred) are from the top-performing teams from each modality (team ‘UZH’ for CT and team ‘KDH’ for MR). Hard situations are highlighted with zoomed-in views in colored borders. In each hard situation, the most illustrative error is marked with a red arrowhead. Case-specific metrics are also reported. Note that the side-road F1 score shown here is computed differently from the main text, as true-negative classes are excluded from the per-class average for single-case evaluation. Abbreviations in metrics: neighbor (Nb) and source (Src).

## 4. Discussions

### 4.1. nnUNet is Necessary but Not Sufficient

All top-3 teams for both modalities used nnUNet, making it a de facto prerequisite for competitive performance. However, nnUNet alone is not sufficient, as many other participating teams also built on nnUNet yet fell significantly short of the top-3. The performance gap between the best and lowest-ranked nnUNet teams can be as large as 40 percentage points in Dice, F1, and contamination scores, highlighting that the choice of framework is far less decisive than how it is used. The top teams maximized the capacity of nnUNet through system design decisions such as pre-training, mixed-modality data, multi-stage pipelines, and post-processing.

### 4.2. Fine-Grained Anatomy Reveals Volume Bias

The more coarse-grained the anatomy, the more large vessels can inflate the overall segmentation scores, obscuring the poor performance on the many smaller and clinically relevant vessels. For instance, when our multiclass predictions are merged into a binary label, the top-3 teams achieved Dice scores of 87–94% for CTA and 93–95% for MRA, without any binary-specific training. However, these high scores are inflated by large vessels that dominate the voxel space and are inherently easier to segment. Our fine-grained anatomical evaluation shows that segmentation difficulty varies substantially across vessel classes: vessels above 3 mm in diameter all achieve Dice scores above 86%, while many smaller vessels lag significantly behind, especially in the four “hard situations”. Fine-grained anatomy therefore makes the volume bias visible, highlighting that small and complex vessels remain the true bottleneck in brain vessel segmentation.

### 4.3. Complementary Contamination Metrics

Standard segmentation metrics such as Dice and Hausdorff distance quantify per-class overlap but do not characterize inter-class prediction errors. The proposed contamination metrics complement standard segmentation metrics by providing a more interpretable breakdown of where and how predictions fail: whether errors are concentrated within foreground regions or scattered across the background, along with the number of contributing classes. The metrics show a good correlation with overall performance — the top-3 teams on the main leaderboard also tend to achieve the best contamination scores. Furthermore, the contamination scores are consistent with the qualitative results shown in Fig. 5, where higher contamination values correspond to visually apparent inter-class confusion and spurious background predictions. To our knowledge, the proposed contamination metrics are the first to explicitly address inter-class confusion in the context of multiclass segmentation.

### 4.4. Robustness to Stroke Pathologies

The TopCoW images used in this challenge were collected from a stroke center, consisting of an elderly patient cohort with a high prevalence of neurovascular pathologies including large vessel occlusion (LVO) and stenosis. The qualitative results in Fig. 5 illustrate this: the MRA case has LVO at the R-ICA and stenosis of the R-VA near the VB junction and at the R-MCA M1-M2 junction. Despite these pathological variations, the top-performing models produce robust segmentations, which is expected given that the training set contains similar stroke and stenosis cases. This suggests that the trained models are already well-suited for deployment in stroke-center settings, where such pathologies are routinely encountered.

### 4.5. Limitations and Future Work

The test set size is a limiting factor for this benchmark, with only 20 MRA and 20 CTA cases, which may affect the stability of the reported metrics. The vessel diameter measurements used in the caliber-performance analysis were derived from the TopCoW patient cohort, which includes cases with stenosis and LVO; as these pathologies directly affect vessel caliber, the reported diameters may not reflect true healthy vessel dimensions and should be interpreted with caution. Nevertheless, the TopBrain dataset and the segmentation models trained in this challenge can serve as a basis for more accurate measurements on other cohorts.

More broadly, the fine-grained vessel segmentation enables two clinically relevant downstream applications. First, the detailed anatomical labeling allows precise localization of pathologies such as LVO, stenosis, and aneurysm on the vascular tree, identifying not just their presence but their exact anatomical location. Second, since stenosis and LVO are fundamentally diseases of vessel caliber change, the caliber analysis workflow demonstrated in this work can be extended to detect and quantify these pathologies directly from vessel diameter measurements derived from the segmentation.

## 5. Conclusion

TopBrain 2025 Challenge introduced the first benchmark for multiclass segmentation of 48 landmark brain vessels for both arterial and venous systems in CT and MR angiography. We demonstrated that fine-grained anatomical definitions reveal segmentation difficulties that coarser benchmarks obscure, and that novel contamination metrics offer complementary insights beyond standard overlap measures. The top-performing teams showed that nnUNet, when combined with principled system-level design choices, can achieve strong performance on this challenging task. Although room for improvement remains, particularly for small and complex vessels.

We believe accurate and fine-grained brain vessel segmentation helps development of automated tools for stroke diagnosis, aneurysm localization, and stenosis quantification. We hope TopBrain has demonstrated the feasibility of this vision and inspired the community to push towards its clinical realization.

## Appendix A. Anatomy abbreviations

The anatomy abbreviations used in this paper are listed below in their vessel regions alphabetically:

### Vertebrobasilar (VB)

**AICA**: anterior inferior cerebellar artery; **BA**: basilar artery; **PICA**: posterior inferior cerebellar artery; **PPTA**: persistent primitive trigeminal artery; **SCA**: superior cerebellar artery; **VA**: vertebral artery.

### Posterior cerebral artery (PCA)

**CalA**: calcarine artery; **P1, P2, P3, P4**: P1 to P4 segments of the PCA; **PaOcA**: parieto-occipital artery; **QC**: quadrigeminal cistern.

### Internal carotid artery (ICA)

**AChA**: anterior choroidal artery; **DDR**: distal dural ring; **OA**: ophthalmic artery; **Pcom**: posterior communicating artery.

### Anterior cerebral artery (ACA)

**A1, A2, A3**: A1 to A3 segments of the ACA; **Acom**: anterior communicating artery; **CC**: corpus callosum; **CMA**: callosomarginal artery.

### Middle cerebral artery (MCA)

**Insula**: insular cortex; **M1, M2, M3**: M1 to M3 segments of the MCA; **Oper**: operculum; **Sphe**: sphenoidal compartment of Sylvian fissure.

### External carotid artery (ECA)

**MaxA**: maxillary artery; **MMA**: middle meningeal artery; **STA**: superficial temporal artery.

### Vein

**ACV**: anterior cerebral vein; **ASV**: anterior septal vein; **BVR**: basal vein of Rosenthal; **CS**: confluence of sinuses; **DMCV**: deep middle cerebral vein; **ICV**: internal cerebral vein; **ISS**: inferior sagittal sinus; **StS**: straight sinus; **SPS**: superior petrosal sinus; **SSS**: superior sagittal sinus; **Tentorium**: tentorium cerebelli; **TSV**: thalamostriate vein; **VoG**: vein of Galen.

In the paper, the prefixes ‘L-’ and ‘R-’ in front of anatomy labels mean left and right, respectively.

## Appendix B. Side-road Vessel Distribution

We document the prevalence of side-road vessels from our training and test data in Table B.4.

## Appendix C. Descriptions of Submitted Algorithms and Teams

In this section, we summarize the methods and algorithms of all the participating teams ordered alphabetically by team names:

### ARG (ARG-DeepNeuro)

The team consisted of Dahye Lee and Kwanseok Oh. They employed a two-stage framework: Stage-1 is binary vessel segmentation and Stage-2 is multiclass vessel segmentation using the probability map from Stage-1 as an additional input channel. Training data combined both MRA and CTA modalities of the TopBrain data. Dice and cross-entropy (CE) losses were used in 3D nnUNet [30] architecture. Final model was ensembled using five folds after training for 1,000 epochs using Adam optimizer with an initial learning rate of 1e-2 on a single 11 GB GPU. Connected component-based postprocessing were used to remove small false positives. **Their algorithms have been included in our Zenodo Docker release (Zenodo record 20158639)**.

**Table B.4.**
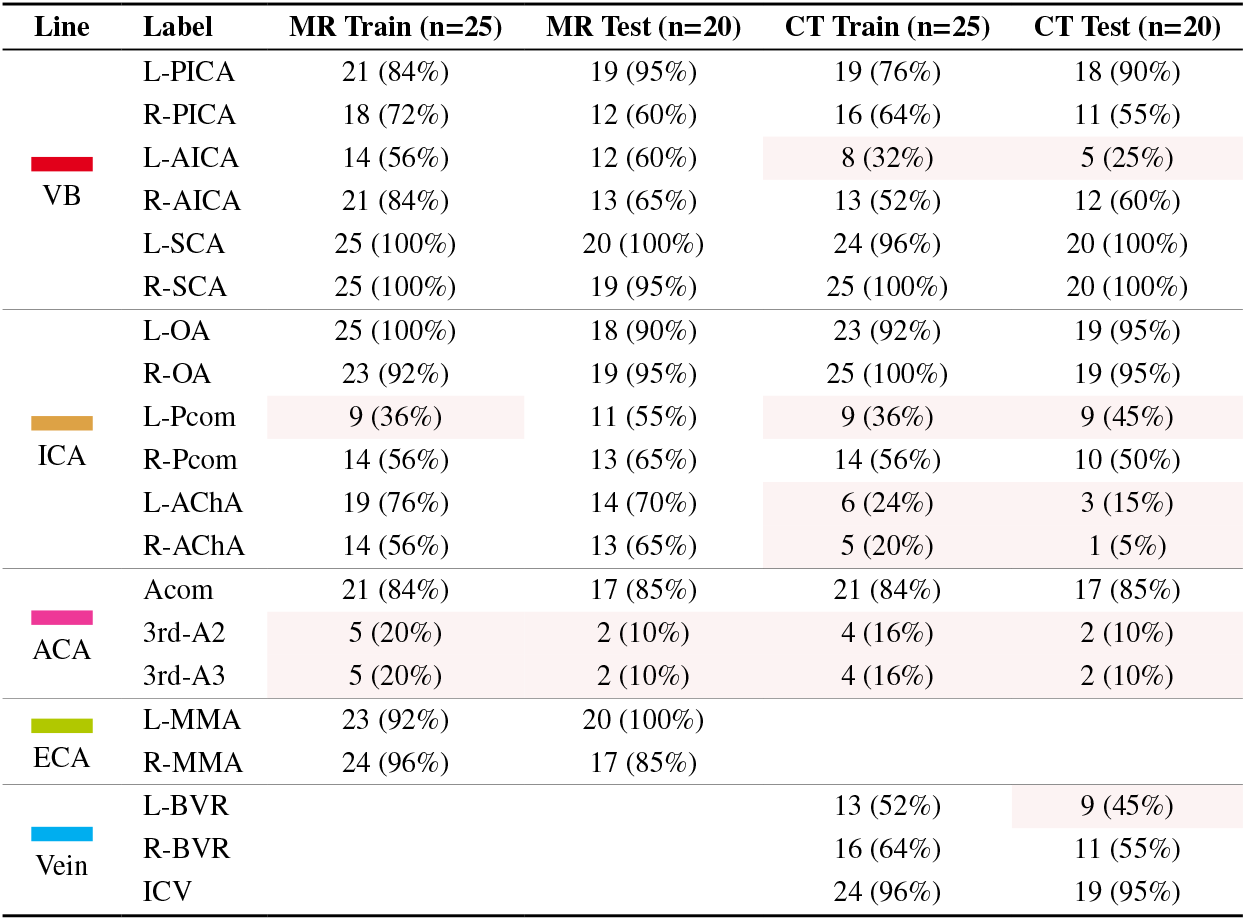
Distribution of side-road vessel presence in training and test sets. Values displayed are count and percentage in brackets. Vessels with percentage below 50% are highlighted in light red.

| Line | Label | MR Train (n=25) | MR Test (n=20) | CT Train (n=25) | CT Test (n=20) |
| --- | --- | --- | --- | --- | --- |
| VB | L-PICA | 21 (84%) | 19 (95%) | 19 (76%) | 18 (90%) |
|  | R-PICA | 18 (72%) | 12 (60%) | 16 (64%) | 11 (55%) |
|  | L-AICA | 14 (56%) | 12 (60%) | 8 (32%) | 5 (25%) |
|  | R-AICA | 21 (84%) | 13 (65%) | 13 (52%) | 12 (60%) |
|  | L-SCA | 25 (100%) | 20 (100%) | 24 (96%) | 20 (100%) |
|  | R-SCA | 25 (100%) | 19 (95%) | 25 (100%) | 20 (100%) |
| ICA | L-OA | 25 (100%) | 18 (90%) | 23 (92%) | 19 (95%) |
|  | R-OA | 23 (92%) | 19 (95%) | 25 (100%) | 19 (95%) |
|  | L-Pcom | 9 (36%) | 11 (55%) | 9 (36%) | 9 (45%) |
|  | R-Pcom | 14 (56%) | 13 (65%) | 14 (56%) | 10 (50%) |
|  | L-AChA | 19 (76%) | 14 (70%) | 6 (24%) | 3 (15%) |
|  | R-AChA | 14 (56%) | 13 (65%) | 5 (20%) | 1 (5%) |
| ACA | Acom | 21 (84%) | 17 (85%) | 21 (84%) | 17 (85%) |
|  | 3rd-A2 | 5 (20%) | 2 (10%) | 4 (16%) | 2 (10%) |
|  | 3rd-A3 | 5 (20%) | 2 (10%) | 4 (16%) | 2 (10%) |
| ECA | L-MMA | 23 (92%) | 20 (100%) |  |  |
|  | R-MMA | 24 (96%) | 17 (85%) |  |  |
| Vein | L-BVR |  |  | 13 (52%) | 9 (45%) |
|  | R-BVR |  |  | 16 (64%) | 11 (55%) |
|  | ICV |  |  | 24 (96%) | 19 (95%) |

### CLAIM

The team was composed of Adam Hilbert and Orhun Utku Aydin. They utilized the ResEnc-L architecture from the nnSSL framework [31], specifically tailored for the MRA modality through a self-supervised learning (SSL) pre-training strategy. The model was pre-trained on a dataset of around 1.7k public MRA images [32] for 1,000 epochs using a single-fold approach and default nnSSL configurations. nnUNet framework was used for fine-tuning, employing a 5-fold cross-validation on the TopBrain MRA dataset and with ‘Deep supervision’ turned off. The prediction was aggregated using majority voting across three selected folds.

### DLaBella29

The submissions were made by Dominic LaBella, who participated in the MRA track. He used the SegResNetDS backbone from the MONAI Auto3DSeg framework [33, 34] with a single MRA channel. The model was trained using AdamW optimizer for 600 epochs on 10 fold cross-validation using DiceCELoss. 5-fold STAPLE [35] fusion was used with no additional pre/post-processing.

### Dolphins

The team was composed of Moona Mazher, Steven A. Niederer, and Abdul Qayyum. The model was trained for the MRA modality using the TopBrain MRA data with an emphasis on full brain vessel context. They designed a custom mix network that incorporates a bidirectional quasi-separable mixing module within a 3D encoder-decoder architecture to capture long-range spatial dependencies. The mix network involved a casual pass with semi-separable state space model (SSM) and shift, an anti-causal pass with reversed sequence and shifts, and diagonal self-connections.

### IMR

The team was composed of Yaoyu Liu, Minghui Zhang, and Yun Gu. The model was trained within the nnUNet frame-work with a custom connectivity-aware loss (CAL) based on prior implementations [36, 37] to improve topological completeness. The total loss included a Dice with focal loss for basic segmentation, and a dynamically weighted loss for tubular multiclass segmentation: for each patch volume, each class of the multiclass vessel was dynamically weighted based on a distance transform to the centerline.

### JQC (junqiangchen)

The submissions were made by Junqiang Chen. He employed a unified architecture based on VNet3D [38], training two modality-specific models independently on the CTA and MRA subsets of the TopBrain data. The images were pre-processed by scaling to the same size after intensity normalization. The model was trained using AdamW optimizer for 5,000 epochs using Dice and CE losses.

### KDH (KPopDemonHunters)

The team consisted of Wooseung Kim, Napasara Asawalertsak, Minjae Kim, Dongho Shin, and Sung-Hong Park. For the MRA track, the model was developed using four strategies. Strategy-1 was using the self-supervised masked autoencoder (MAE) pre-trained network from OpenMind [31, 39]. Strategy-2 was fine-tuning pretrained ResEnc-UNet with around 1,000 public MRA scans for training a nnUNet model. Strategy-3 was using a separate binary segmentation model to remove false positives from the multiclass segmentation output. Strategy-4 was making pseudo-label on the remaining 100 TopCoW MRA scans [7]. The MRA model was ensembled after training for five fold cross-validation using the skeleton recall (SkelRecall) loss. For the CTA track, the model was trained after 1,000 epochs employing only Strategy-4 of pseudo-label with TopCoW CTA data and without SkelRecall loss. **Their algorithms have been included in our Zenodo Docker release (Zenodo record 20158639)**.

### negichi

The team was composed of Shunsuke Kikuchi. The models were trained for 4,000 epochs for MRA and 3,500 epochs for CTA using the default 3D nnUNet in full-resolution mode.

### refrain

The submissions were made by Yaqing Zhang, Jialu Liu, and Yue Cui. They employed a MR/CT modality harmonization preprocessing that involved intensity truncation, universal resample, and normalization. They also performed offline data augmentation for subjects with rarer vessel classes. Based on anatomical regions, the vessel classes were divided into five groups, and five models were trained accordingly. Three models were trained and inferred after region-of-interest (ROI) extraction, which was done by a standard-space ROI template registered to individual space for cropping. They used 3D Res-UNet as the backbone with default nnUNet settings and trained the models for 200 epochs. The predictions from the five models were merged and resampled back to the original resolution.

### U&E

The team comprised Yuchen Qiu, Anouk Verschuur, Jiaxin Zhang, Irene van der Schaaf, Ruisheng Su, and Chantal Tax. They adopted a two-stage segmentation strategy built on the nnUNet framework. Stage-1 was a binary vessel segmentation model that served as a vessel prior. Stage-2 was a multiclass vessel segmentation model trained by concatenating the vessel probability map from Stage-1 with the original image as an additional input channel. The MRA and CTA models were separately trained using the respective modality from the TopBrain data. For the MRA model, the Stage-1 network was initialized with pretrained weights from an in-house vessel–aneurysm segmentation model. The CTA model did not have any pretrained initialization. The models were trained using the 3D full-resolution mode of nnUNet for 1,000 epochs with Dice, CE, and TopK losses. To reduce memory usage, a chunk-based sliding-window strategy was implemented.

### UTRad

The team consisted of Yosuke Yamagishi, Nishta Letchumanan, and Shouhei Hanaoka. They fine-tuned a vessel foundation model, vesselFM, based on the DynUNet architecture [40]. Preprocessing included isotropic resampling, percentile normalization, random cropping of 96×96×96 cubes. The CTA model was trained for 500 epochs, while the MRA model was trained for 1,000 epochs, using the DiceFocalLoss. Inference was performed with sliding window of the crop-sized cubes.

### UZH

The team was composed of Houjing Huang, Kaiyuan Yang, and Pengcheng Shi. Vessel classes were grouped into anatomical regions, yielding five models (two folds each) trained on both modalities where applicable. Inter-modal registration for all the training image pairs was employed as a data augmentation strategy. All models were based on the nnUNet framework and mirrored the architecture of the 2024 TopCoW UZH submission, with TopK loss newly integrated into the loss function. The original TopCoW model weights were used for training initialization. Postprocessing involved the removal of small components below a threshold of 12 voxels. **Their algorithms have been included in our Zenodo Docker release (Zenodo record 20158639)**.

### VdHi

The team was composed of Jesús González and Riccardo Tiberi. There were two stages to the segmentation model: Stage-1 was a binary vessel segmentation, and Stage-2 was the multiclass vessel segmentation. Both stages used nnUNet with the full-resolution 3D configuration. Preprocessing included bilateral filtering to denoise while preserving vessel edges. The predictions were fused via seeded watershed, where binary masks constrained the vascular tree and labeled predictions seeded vessel identity for region growing. Postprocessing used an adjacency matrix that encoded relationships between vessel classes to enforce anatomical plausibility. Small islands below 20 voxels were removed, and labels were corrected using the consistency rules.

### ViCOROB

The team comprised Rachika E. Hamadache, Clara Lisazo, Cansu Yalcin, Valeriia Abramova, Uma M. Lal-Trehan Estrada, Agustin Cartaya Lathulerie, Micaela Rivas Díaz, Adrià Casamitjana, Arnau Oliver, and Xavier Lladó. The pipeline had three stages of segmentation, trained using 3D full-resolution nnUNet with Dice, CE, and centerline boundary Dice (cbDice) [41] losses. Stage-1 was binary vessel segmentation. Stage-2 was intermediate multiclass segmentation trained by clustering the vessel classes to 14 coarse-grained labels based on anatomical regions. Stage-3 was full multiclass segmentation. The final prediction was generated by combining the outputs from all stages. Postprocessing included iterative background filling and small components removal using the binary segmentation mask.

### “6100”

The team consisted of Menghan Zhang and Zhiqiang Bai. The segmentation model was based on the nnUNet framework, with a multi-scale pyramid fusion (MSPF) module integrated into the skip connections. The model was trained on the MRA dataset provided by the challenge, using the standard nnUNet preprocessing and data augmentation pipelines.

## Data Availability

The TopBrain training data of 50 annotated volumes is released in our public Zenodo repository at https://zenodo.org/records/16878417.

## Code Availability

The Docker images from best performing teams and the scripts to help run them locally are released in our public Zenodo repository at https://zenodo.org/records/20158639.

The implementation of our evaluation metric code is open sourced at https://github.com/CoWBenchmark/TopBrain_Eval_Metrics.

## Acknowledgments

The TopBrain Challenge is supported by the Helmut Horten Foundation. FM, NJ, and SH received funding from the European Union’s Horizon Europe research and innovation programme under grant agreement No 101136438 (GEMINI project).

Team **ARG** (ARG-DeepNeuro) was supported by the Technology Innovation Program (or Industrial Strategic Technology Development Program) (RS-2025-02221011, Development of Medical-Specialized Multimodal Hyperscale Generative AI Technology for Global Integration) funded by the Ministry of Trade, Industry & Energy (MOTIE, Korea) and the Institute of Information & Communications Technology Planning & Evaluation (IITP) grant funded by the Korea government (MSIT) No. RS-2022-II220959 ((Part 2) Few-Shot Learning of Causal Inference in Vision and Language for Decision Making). Team **U&E**’s CMWT is supported by the Dutch Research Council (NWO) under a VIDI grant with file number 21299.

## Footnotes

^2^Data at https://zenodo.org/records/16878417 and Dockers at https://zenodo.org/records/20158639

